# Simultaneous evaluation of antibodies that inhibit SARS-CoV-2 RBD variants with a novel competitive multiplex assay

**DOI:** 10.1101/2021.03.20.21254037

**Authors:** Ester Lopez, Ebene R. Haycroft, Amy Adair, Francesca L. Mordant, Matthew T. O’Neill, Phillip Pymm, Samuel Redmond, Nicholas A. Gherardin, Adam K. Wheatley, Jennifer. A. Juno, Kevin J. Selva, Samantha Davis, Leigh Harty, Damian F.J. Purcell, Kanta Subbarao, Dale I. Godfrey, Stephen J. Kent, Wai-Hong Tham, Amy W. Chung

**Author notes:** **Correspondence:** Amy W Chung, Department of Microbiology and Immunology, Peter Doherty Institute for Infection and Immunity, The University of Melbourne, Victoria 3000, Australia.

## Abstract

The SARS-CoV-2 Receptor Binding Domain (RBD) is both the principal target of neutralizing antibodies, and one of the most rapidly evolving domains, which can result in the emergence of immune escape mutations limiting the effectiveness of vaccines and antibody therapeutics. To facilitate surveillance, we developed a rapid, high-throughput, multiplex assay able to assess the inhibitory response of antibodies to 24 RBD natural variants simultaneously. We demonstrate that immune escape can occur through two mechanisms, antibodies that fail to recognize mutations, along with antibodies that have reduced inhibitory capacity due to enhanced variant RBD-ACE2 affinity. A competitive approach where antibodies simultaneously compete with ACE2 for binding to the RBD may therefore more accurately reflect the physiological dynamics of infection. We describe the enhanced affinity of RBD variants N439K, S477N, Q493L, S494P and N501Y to the ACE2 receptor, and demonstrate the ability of this assay to bridge a major gap for SARS-CoV-2 research; informing selection of complementary monoclonal antibody candidates and the rapid identification of immune escape to emerging RBD variants following vaccination or natural infection.

## INTRODUCTION

Severe acute respiratory syndrome coronavirus 2 (SARS-CoV-2) was first identified in late 2019 in Wuhan, China, (1, 2) and has since become an ongoing global public health emergency. Coronavirus disease (COVID-19) is the clinical syndrome associated with SARS-CoV-2 infection, which is characterized by a respiratory syndrome with a variable degree of severity. To date millions deaths have been reported (3), with the current pandemic not only threatening public health, but also adversely impacting economies worldwide. As we enter the second year of the SARS-CoV-2 pandemic, and promising vaccines are rolled out (4, 5), the ongoing transmission along with the emergence of new variants with multiple mutations at key residues in the spike glycoprotein (6-8), has resulted in the continued implementation of stringent public health measures to control infection in many parts of the world.

SARS-CoV-2 enters host cells via the angiotensin-converting enzyme 2 (ACE2) receptor (9-11). Binding to ACE2 and entry to the host cell is facilitated via the “ spike”, a homo-trimeric transmembrane envelope glycoprotein which consists of the binding (S1), and transmembrane-fusion (S2) domains that are processed from the polyprotein precursor at a polybasic furin-cleavage site (9, 12). Key to this virus-host interaction is the receptor-binding domain (RBD), which lies within the S1 subunit, and is critical for binding to ACE2 receptors on target cells (11). Numerous studies have demonstrated that potent neutralizing antibodies (NAbs) that recognize the RBD are consistently elicited following infection (13-16), and account for most plasma neutralizing activity, and inhibition of RBD-ACE2 binding (13, 17). Antibodies targeting the RBD account for ∼90% of the neutralizing activity present in SARS-CoV-2 immune sera (17), and therefore have great potential to be clinically useful in the treatment and prevention of SARS-CoV-2 infection. The RBD of SARS-CoV-2 therefore represents a key target for the development of vaccine elicited humoral immunity, and eliciting potent neutralizing antibodies capable of blocking ACE2 binding remains a key feature of the development of effective vaccines and antibody therapies.

As for any viral outbreak, of great concern is the emergence of gain of function variants which facilitate viral infectivity, transmissibility, or neutralizing antibody escape. Since the beginning of the pandemic, SARS-CoV-2 genomic sequencing has identified a number of variants revealing a modest rate of evolution of the viral genome (18, 19). This information has been made available through public repositories such as the Global Initiative on Sharing All Influenza Data (GISAID) (20) which have been critical to monitoring the epidemiology of the virus and informing SARS-CoV-2 research. Considering the critical role of the RBD in mediating viral entry, the tendency of the RBD of SARS-CoV coronaviruses to be highly variable (21), and the recent emergence of new variants B.1.1.7, P.1 and B.1.35 (6-8, 22), increased interest has been directed toward the surveillance of SARS-CoV-2 RBD mutations. Of concern is the observation that several of these SARS-CoV-2 RBD mutations escape monoclonal antibody (mAb) neutralization and/or attenuate polyclonal plasma neutralization (23-27). Subsequently, there has been a more cautious shift in assessing the efficacy of current vaccine and antibody therapies with respect to the continuously emerging genetic variants, especially since some vaccine candidates and antibody therapies solely target the wild-type RBD (28, 29).

Though there is no standardized method for assessing the antibody-mediated neutralization of SARS-CoV-2, which can account for variation in between studies (30). Assays which evaluate the ability of NAbs to inhibit viral replication in target cells by observing plaque reduction or a cytopathogenic effect are currently regarded as benchmark assays. The pseudovirus-based neutralization assays (31) which reduce the biosafety requirements when working with SARS-CoV-2 virus can also be used, however these cell culture based assays can be challenging to implement and time-consuming to run, which limits scalability. Until recently these were the only conventional assays available for evaluating NAbs. Limitations of the aforementioned assays have however motivated efforts to develop alternative assays such as the surrogate virus neutralization test (sVNT) (32) which mirrors the RBD-ACE2 interaction in an ELISA plate format, a novel fluorescence reporter cell-based assay (33), and lentiviral particle fluorescent neutralization assays (23, 34). These assays are however limited to screening NAbs directed to only one RBD mutant at a time. In addition, these assays allow antibodies to incubate and neutralize SARS-CoV-2 before incubation with soluble ACE2 or ACE2 expressing cells. This approach however does not consider that physiologically antibodies may have to compete with ACE2 for binding to the RBD, and that some of these RBD variants have a significantly enhanced affinity for ACE2. The absence of a competitive assay to evaluate the neutralizing potential of SARS-CoV-2 antibodies to multiple emerging RBD variants simultaneously thus presents a major gap for SARS-CoV-2 surveillance, and effective vaccine and antibody therapy development.

Herein we describe the development of a novel high-throughput RBD-ACE2 multiplex inhibition assay that measures SARS-CoV-2 NAbs to multiple RBD natural variants simultaneously. Multiplex assays provide several key advantages including less sample volume requirements, and the ability to reliably detect analytes across a broad dynamic range to provide a much higher resolution of data than conventional immunoassays like ELISA (35, 36). We describe the validation of this rapid, high-throughput multiplex assay, and evaluate how a selection of naturally occurring single amino acid RBD variants observed during viral surveillance impact the potential efficacy of mAb therapeutics and the recognition by polyclonal convalescent human plasma. Our competitive neutralization multiplex assay allows antibodies to simultaneously compete with host cell receptor ACE2 for binding to an array of RBD variants coupled to magnetic beads, an approach which better captures the physiological dynamics of the interaction, which we demonstrate provides an enhanced resolution of the SARS-CoV-2 RBD-ACE2 NAb response. We also reveal how this multiplex assay can serve to flag RBD variants which may have an enhanced affinity for ACE2, describe and compare the binding kinetics of several of these enhanced affinity RBD variants, and demonstrate how this enhanced affinity to ACE2 can reduce the inhibitory capacity of both mAbs and convalescent plasma.

## RESULTS

With the development of alternative ELISA-based assays which mimic the virus-receptor interaction (32, 37), we sought to evaluate whether a similar approach could be adapted to a bead-based multiplex assay format. In this novel approach to evaluate SARS-CoV-2 neutralizing antibodies, recombinant RBD was coupled to magnetic multiplex beads. Convalescent plasma or purified mAbs are then allowed to simultaneously compete with soluble biotin-ACE2 for binding to the RBD coupled onto the magnetic microspheres Figure 1A. This approach was first validated with wild-type (WT; Wuhan strain) RBD and Spike 1 (S1) of SARS-CoV-2 to demonstrate proof of concept, before subsequent development of a multiplexed SARS-CoV-2 natural RBD variant array.

**Figure 1.**
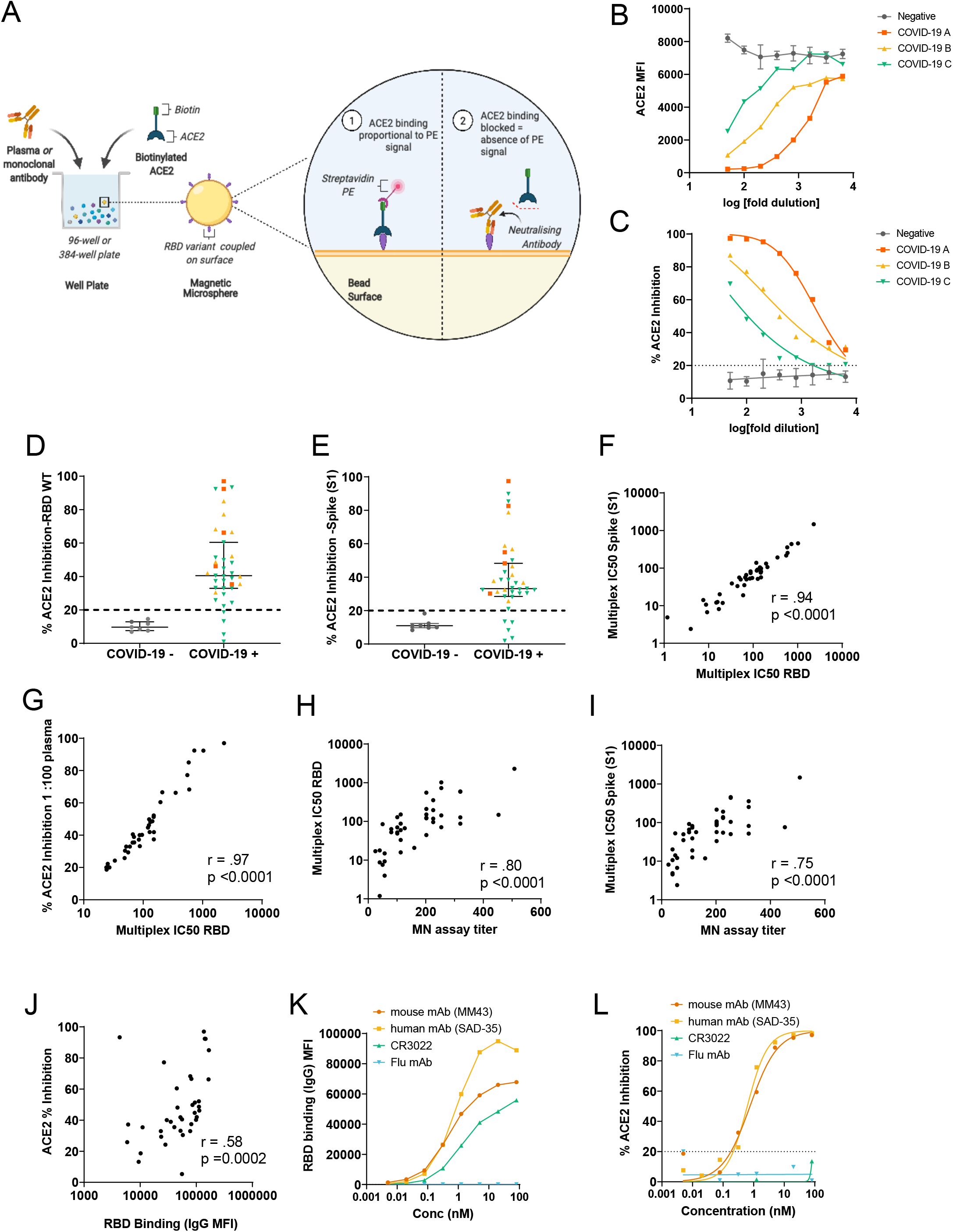
Principle and validation of the SARS-CoV-2 inhibition assay. **A**. Schematic representation of the RBD multiplex assay: Anti-SARS-CoV-2 RBD antibodies compete with biotin-conjugated ACE2 for binding to RBD variants coupled to magnetic microspheres. **B**. Inhibition of the RBD-ACE2 interaction by COVID-19 subjects recovered from SARS-CoV-2 infection (COVID-19 A, B and C) vs Mean ±S.D of (*n*=7) COVID-19 negative samples, with a decrease in MFI observed as anti-SARS-CoV-2 neutralizing antibodies present in the sample block the ACE2 binding RBD coupled to magnetic microspheres. **C**. Inhibition of the SARS-CoV-2 RBD–ACE2 interaction by plasma from convalescent COVID-19 subjects, with the determined cut-off at 20% inhibition indicated by the dotted line. Illness severity was severe for example COVID-19 subject A (orange-squares) moderate for B (yellow-triangles) and mild for subject C (green-inverted triangles). Illustrations were created using BioRender. **D**. Plasma samples from subjects recovered from SARS-CoV-2 infection (*n* = 37) and SARS-CoV-2 negative healthy controls (*n* = 7) were tested for inhibition of the RBD-ACE2 interaction and **E**. the Spike (S1)-ACE2 interaction. SARS-CoV-2 positive subjects are colored based on COVID-19 illness severity. Orange Squares=Severe, Yellow Triangles=Moderate, Green Inverted Triangles=Mild. Grey = SARS-CoV-2 negative controls. **F**. Spearman’ s Correlation between relative Spike (S1) and RBD IC_50_. percentage inhibition values. **G**. Spearman’ s Correlation of the percentage ACE2 inhibition at a 1 in 100 dilution of plasma to RBD and the relative IC_50_. value. **H**. Spearman’ s Correlation of Multiplex ACE-RBD inhibition assay IC_50_ values and the titer obtained with the virus microneutralization assay. **I**. Spearman’ s Correlation of Multiplex ACE-Spike (S1) inhibition assay IC_50_.values and the titer obtained with the virus microneutralization assay. **J**. Spearman’ s Correlation of ACE-RBD inhibition and RBD Binding **K**. RBD binding of control mAbs **L**. Percentage ACE2 inhibition of control mAbs.

### Validation of a novel multiplex RBD-ACE2 inhibition assay to measure SARS-CoV-2 neutralizing antibodies in plasma to WT RBD and Spike (S1)

A subset of previously described (38) plasma samples collected from a cross-sectional cohort of Australian adults recovered from SARS-CoV-2 infection were chosen to firstly evaluate the performance of this assay with regard to its ability to quantitate ACE2 inhibitory antibodies from SARS-CoV-2 convalescent subjects. Subsequently, we then sought to determine whether NAbs in the SARS-CoV-2 positive plasma could inhibit ACE2 receptor binding in a similar manner as the virus microneutralization assay (39). In this assay, inhibition of the SARS-CoV-2 RBD-ACE2 interaction by antibodies in SARS-CoV-2 convalescent plasma was observed as dose-dependent decrease in fluorescent ACE2 measured as MFI (Median Fluorescence Intensity) bound to immobilised RBD, whereas the MFI of SARS-CoV-2 negative plasma remained high across the 8-point 2-fold serial dilution (Figure 1B), demonstrating that the specific inhibition of the SARS-CoV-2 RBD–ACE2 interaction by plasma from convalescent SARS-CoV-2 subjects occurs in a dose-dependent manner (Figure 1C). The capacity of antibodies in SARS-CoV-2 convalescent plasma to inhibit the interaction between ACE2 and RBD in this assay was detected in most subjects in this cohort (median, 40.5%; IQR, 32.9%–60.4%), with 31% subjects exhibiting more than 50% ACE2 inhibition activity Figure 1D. In comparison, just under a quarter (23%) of subjects exhibited more than 50% inhibition activity against S1 (median, 33.1%; IQR, 28.4%–48.3%) (Figure 1E), suggesting that the full S1 protein may provide additional stability to the ACE2-RBD interaction. A nominal cut-off of 20% was set based on a panel of SARS-CoV-2 negative subjects (Supplementary Figure 1A. To further validate this assay, each plasma sample’ s relative half-maximum inhibitory concentration (IC_50_) titer to RBD (WT) and spike (S1) was determined from the 2-fold 8-point dilution. Relative IC_50_values of the ACE2-RBD versus ACE2-S1 interaction by plasma NAbs strongly correlated (r=.94, p< 0.0001) Figure 1F, and calculated ACE2 % inhibition at a 1 in 100 dilution of plasma positively correlated with RBD IC_50_, (r=.96, p< 0.0001) Figure 1G, suggesting that a 1 in 100 dilution of plasma would be closely representative of the relative IC_50_.

In order to determine how well this assay could detect NAbs in comparison to a virus neutralization assay (39), relative IC_50_ values to RBD and S1 were correlated to a microneutralization assay using SARS-CoV-2 infection of Vero cells. As shown in Figure 1H and Figure 1I, there was a strong correlation between the two assays for RBD-ACE2 inhibition (r = 0.8, p< 0.0001) and S1-ACE2 inhibition (r= 0.75, p< 0.0001). In addition, we examined the relationship between ACE2 inhibition and IgG RBD binding (Figure 1J), and found a moderate correlation between the two (r = 0.58, p=0.0002), which suggests that inhibition may also be mediated by other antibody isotypes such as IgA and IgM or to other key regions of the S1 protein, such as the N-terminal domain (NTD) (40). The multiplex assay was determined to be robust (r^2^ = 0.9) Supplementary Figure 1B by comparing the repeatability of the % ACE2-RBD inhibition values obtained by the cohort of plasma samples when tested by two operators in two independent experiments on separate days, with the mean % ACE2-RBD inhibition once again demonstrated to correlate strongly to the microneutralization assay (Supplementary Figure 1C).

Considering mAbs are of increasing interest for therapy or prophylaxis of SARS-CoV-2, we evaluated the ability of this assay to specifically measure mAbs that neutralize the ACE2-RBD interaction. Here we tested two commercially available neutralizing antibodies (one human and one mouse), and two negative controls for RBD binding and ACE2 inhibition. The first negative control was an unrelated human mAb against influenza, and the second was CR3022, a neutralizing mAb which has been described to bind RBD at an epitope that does not overlap with the ACE2 binding site of SARS-CoV-2 and therefore is unable to block the ACE2-RBD interaction (41). As demonstrated in Figure 1K and 1L, both the human and mouse neutralizing mAbs demonstrated a very similar pattern of RBD binding and ACE2 inhibition. In contrast the absence of binding and inhibition of ACE2 by a non-specific influenza mAb confirmed the specificity of this assay. CR3022 binding to RBD coincided with the absence of ACE2 inhibition further validating the specific ability of this assay to evaluate neutralizing antibodies which specifically inhibit the RBD-ACE2 receptor interaction.

### Validation of an RBD natural variant multiplex array

Upon validation of the multiplex RBD-ACE2 assay to WT RBD and S1, we sought to expand the assay multiplexing 23 RBD natural mutants selected from the GISAID RBD surveillance repository at the time (June 2020), as well as RBD variant S477N which emerged and rose to be the second most frequent variant in the following months. A total 25 variants including the WT were studied. Figure 2A illustrates the position of these variants on the RBD, and Figure 2B displays the more recent observed frequency of these variants according to the GISAID repository, with variant N501Y, currently the most frequent RBD variant worldwide (a key mutation present in the newly emergent B.1.1.7, B.1.351, B.1.1.70 and P.1 strains). The RBD variant array was set up by coupling each of these 25 variants to an individual “ bead region” corresponding to a unique spectral signature, with coupling efficiency across the RBD variant proteins onto the carboxylated magnetic microspheres verified with an anti-His Tag antibody (Supplementary Figure 2). This was determined by comparing the mean MFI of each of the coupled RBD proteins to the WT, with coupling across all the RBD variants determined to be comparable to the WT RBD. We also correlated the IgG binding (r^2^ =0.97) and ACE2 inhibition (r^2^ =0.99) of each of the variants on their own (single-plex) as well as when in combination with all other variants (multi-plexed) (Supplementary Figure 3A and 3B), which confirmed the absence of any undesirable effect or significant loss of signal as a result of multiplexing all RBD variant proteins simultaneously in a single well.

**Figure 2.**
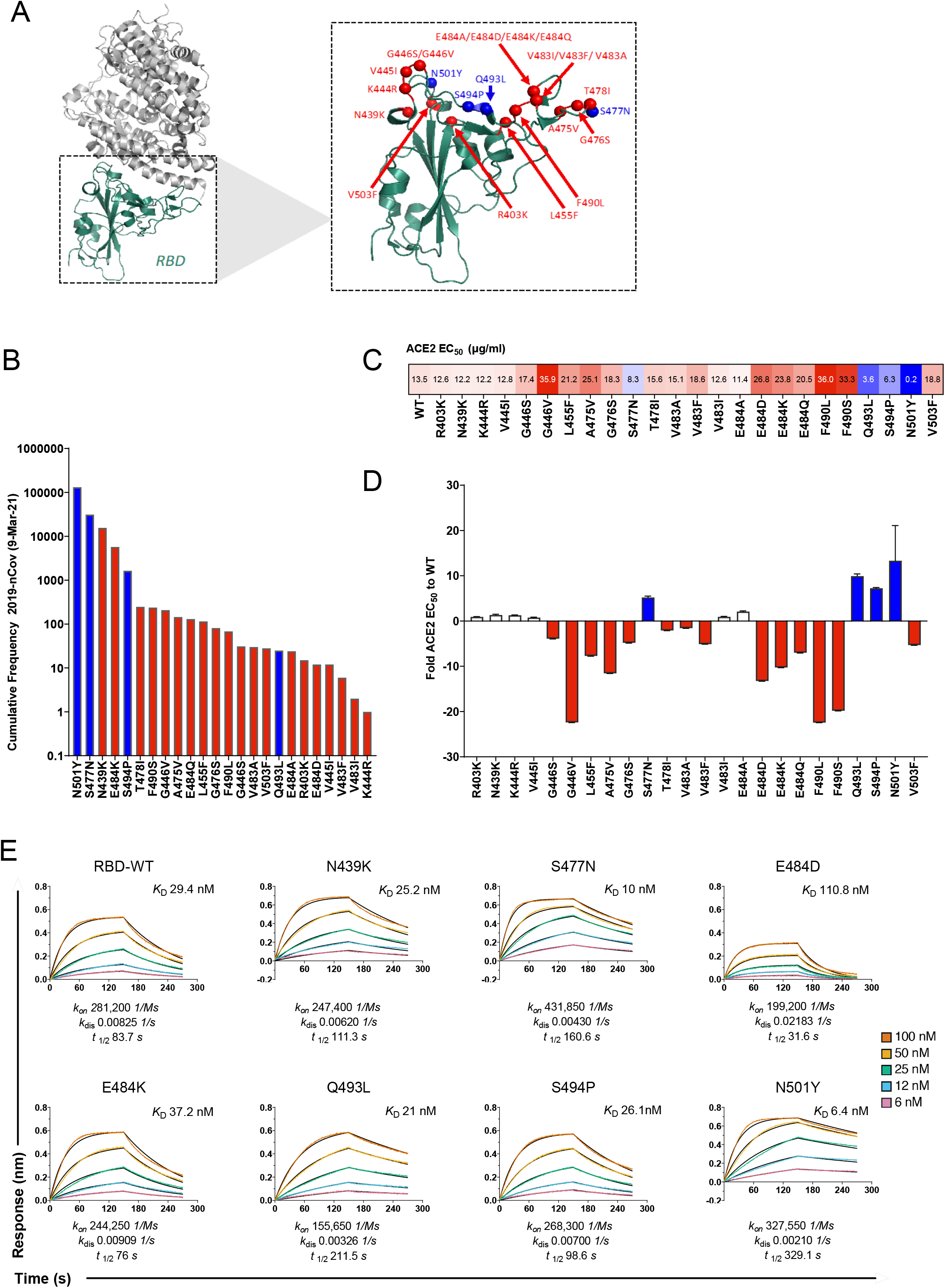
Characterization of RBD variants in the Multiplex array. **A**. Structure of novel coronavirus spike receptor-binding domain (RBD) complexed with its receptor ACE2 (PDB ID code 6LZG) highlighting the amino acid positions of 24 RBD natural variants selected from the GISAID repository in June 2020, with the subsequent inclusion of S477N and their proximity to the RBD-ACE2 interface. The structural illustration was generated using PyMol. Variants with a significantly higher affinity to ACE2 as determined in our multiplex assay are highlighted in blue. **B**. Cumulative Frequency of variants ranked from most frequently isolated to less frequent which are included in the multiplex array. Frequency of genomes is based on high quality 2019-nCov genome sequences reported by the GISAID database as of 9th^th^ March 2021. Variants with a significantly higher affinity to ACE2 as determined in our multiplex assay are highlighted in blue. **C**. Relative EC_50_.values (μg/mL) obtained for each of the variants. Values are color-coded blue for EC_50_ <10 μg/mL and red for EC_50_.values >10 μg/mL. **D**. Mean fold difference for each RBD variant EC_50_ value relative to WT RBD. Data is shown as the mean ± SEM of (*n*=4) independent experiments. Bars for variants with more than a 2-fold difference in affinity to the WT RBD are colored blue. Bars for variants with a slightly increased affinity (< 2-fold) are colored white. Variants with a weaker EC_50_ value relative to WT are coloured red. **E**. Bio-layer interferometry sensograms of immobilized ACE2 with 2-fold 6-100 nM serial dilutions of SARS-CoV-2 RBD variants in solution. Binding curves representative of two independent experiments for each variant are plotted (solid-colored lines), globally fitted to a 1:1 binding model (black line).

**Figure 3.**
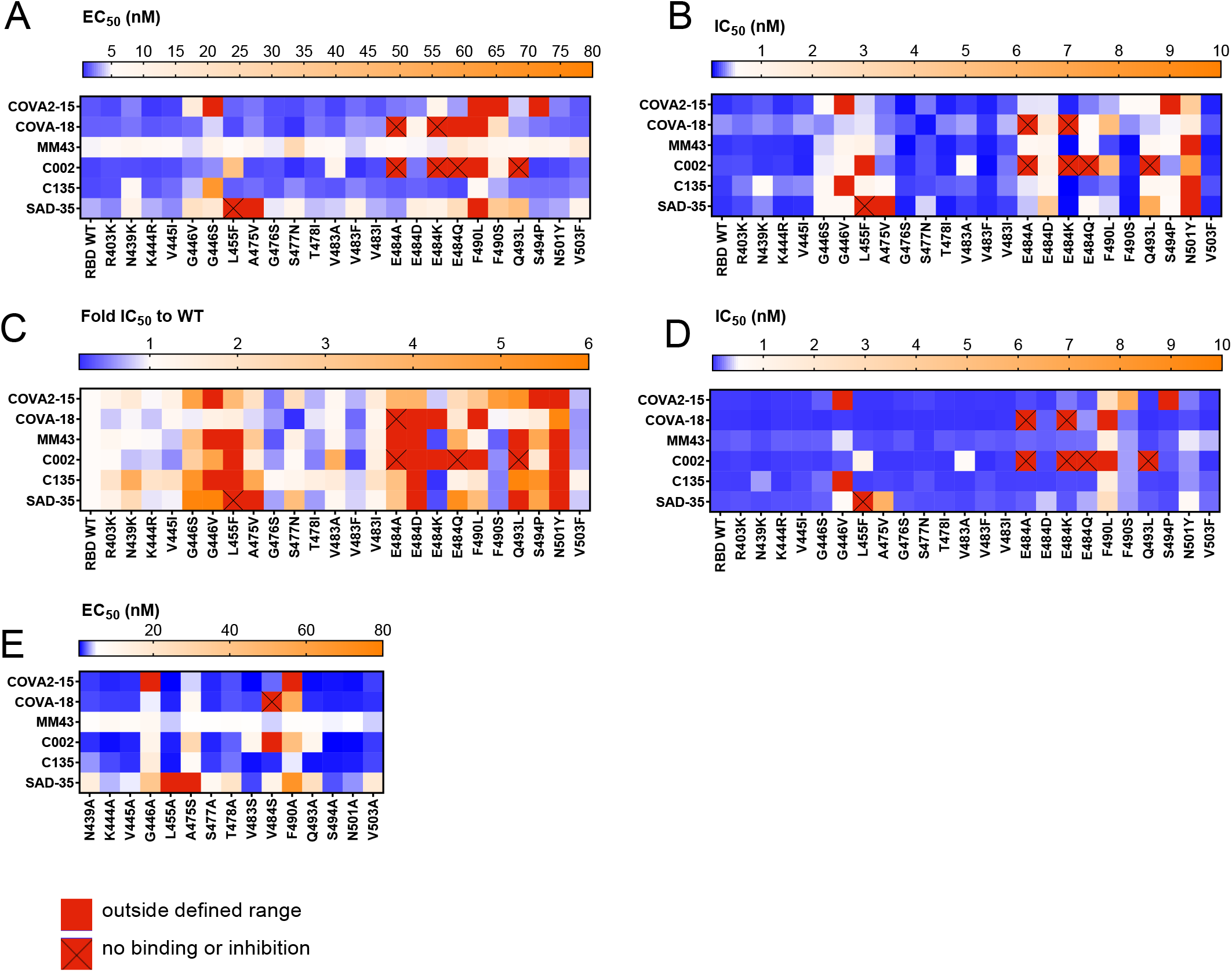
Mapping mutations to the RBD that affect recognition by monoclonal antibodies. **A**. Relative EC_50_. RBD binding of each mAb to each of the RBD variants **B**. Relative IC_50_. RBD inhibition of each mAb to each of the RBD variants. **C**. RBD natural variant ACE2-RBD relative IC_50_. inhibition relative to RBD WT. Blue -Stronger inhibition of RBD variants relative to WT (IC_50_. < RBD WT) Orange-Weaker inhibition of RBD variants relative to WT (>1-6 fold RBD WT IC_50_.). Red >6 fold weaker RBD WT IC_50_.ie. very poor/absence of inhibition. **D**. Relative IC_50_. RBD inhibition of each mAb to each of the RBD variants performed in a non-competitive format where mAbs were pre-incubated prior to addition of ACE2. **E**. Relative EC_50_. Binding of mAbs to Alanine (or Serine if previously alanine) RBD mutants generated of selected RBD variants via multiplex array.

### Affinity of RBD variants to ACE2

To evaluate how the array of RBD variants affected binding to ACE2 in our assay, a 2-fold 12-point serial dilution of ACE2 starting from a final concentration of 160µg/ml per well was performed, and a relative EC_50_ for each of the variants was determined (Figure 2C). The calculated difference in the relative EC_50_ of each variant was then compared to RBD WT (Figure 2D), with the majority of RBD variants exhibiting a similar or weaker relative EC_50_ when compared to the WT RBD. A trend consistent with the observation that many missense mutations often negatively impact ACE2 binding (42). Nevertheless, we found some variants demonstrated significantly enhanced binding to ACE2 relative to WT SARS-CoV-2 RBD. These were N501Y, Q493L, S494P and S477N which demonstrated the lowest overall relative EC_50_ values (0.2, 3.6, 6.3 and 8.3 µ g/ml respectively) as determined by our ACE2-RBD multiplex assay (Figure 2C).

Though it is now well known that N501Y, which is the shared mutation in the B.1.1.7, B.1.351, B.1.1.70 and P.1 lineages, is predicted to have a higher affinity for ACE2, (42), here we sought to characterize the binding kinetics of N501Y to WT RBD and other potential high affinity variants. Based on the relative EC_50_. values observed in our multiplex assay, we selected these high affinity variants, and the emerging N439K variant present in currently circulating linage B.1.258, as well as E484K which is a mutation shared by B.1.351, B.1.525 and P lineages to be profiled and compared for their binding kinetics to ACE2 using Bio-layer interferometry (BLI). In addition, one variant that showed a reduced affinity for ACE2 (E484D), was also selected to be profiled for comparison.

BLI profiles of the RBD variants binding to ACE2 (Figure 2E) confirm that indeed variant N501Y demonstrates an enhanced affinity for the ACE2 receptor, with an almost a 5-fold increase in affinity (*K*_D_ 6.4nM vs 29.4nM), which is driven by a much slower off-rate of this variant (t½ 329.1 s (k_dis_ 0.00210 1/s) vs t½ 83.7 s (k_dis_ 0.00825 1/s)) compared to the WT. Variant S477N also demonstrated enhanced (*K*_D_ 10 nM) affinity for ACE2, although variant Q483L (*K*_D_ 21 nM) demonstrated a slower off-rate t½ 211.5 s vs t½ 160.6 s by comparison. N439K and S494P had weakly enhanced overall affinity for ACE2, *K*_D_ 25.2 nM and 26.1 nM, respectively, compared to the WT RBD (29.4nM). The enhanced affinity of N439K was the result of the almost 1.5 fold slower off-rate t½ 111.3 s (k_dis_ 0.00620 1/s) vs t½ 83.7 s (k_dis_ 0.00825 1/s) of the WT. Confirming multiplex ACE2 EC_50_ assay observations, variants E484K and E484D demonstrated reduced affinity to ACE2 (*K*_D_ 37.2 nM, and *K*_D_ 110.8 nM respectively).

### Mapping mutations to the SARS-CoV-2 RBD that affect recognition by monoclonal antibodies

We next sought to apply the multiplex assay to map potential escape mutations using a panel of six previously characterized mAbs. These were human mAbs COVA2-15 and COVA-18, (43), C002 and C135 (44), SAD-35 (Acro Biosystems) and mouse mAb MM43 (Sino Biologicals). First, we assessed binding of each mAb to the RBD variants by determining the relative EC_50_ (Figure 3A), and second, we determined ACE2 inhibition of these mAbs to these variants by calculating the relative IC_50_ (Figure 3B), from an 8-point 4-fold serial dilution of each mAb. Though all six mAbs bound (Figure 3A) and inhibited (Figure 3B) the SARS-CoV-2 RBD WT with relatively high affinity, they however differed in the extent to which they bound and inhibited each of the RBD variants. Variants with mutations at position 446 and 484 (with the exception of E484D) of the RBD prominently demonstrated antibody escape and consequently poor inhibition. COVA2-15 and C135 demonstrated escape to G446V, while COVA2-15 demonstrated a loss of binding and inhibition to S494P. mAb C002 demonstrated escape from variants E484A, E484K, E484Q and Q493L. Furthermore, N501Y demonstrated an overall reduction in its ability to be inhibited across all mAbs as demonstrated by significantly weaker relative IC_50_ values, compared to the WT (Figure 3C), despite all mAbs having the capacity to bind RBD variant N501Y with relatively high affinity (Figure 3A).

We next generated a panel of 15 of the RBD variants where we mutated the amino acid variants to alanine, (or to serine if alanine was already present) (Figure 3E), in order to further confirm which amino acid positions are essential for mAb epitope binding. We found that for a subset of RBD variants, immune escape occurs due to a reduction or absence of mAb binding at key amino acids, such as position 446 and 490. In contrast, other positions, such 493, 494, and 501 were relatively unaffected by the alanine substitution, suggesting that alternative immune escape mechanisms beyond loss of mAb recognition may be involved, with the attenuation of the neutralizing response, and escape particularly by the N501Y mutation most likely a result of this variant’ s significantly enhanced affinity to ACE2.

The majority of current *in vitro* SARS-CoV-2 neutralization assays first allow antibodies to bind the RBD before subsequent incubation with the ACE2 receptor or ACE2 expressing cells. This approach has the potential to bias the assay to allow for maximal NAb neutralization, thus reducing the influence of RBD variant affinity to the ACE2 receptor. In light of this, we sought to evaluate how mAb pre-incubation followed by incubation with the ACE2 receptor compared to our original competitive format of the ACE2-RBD inhibition assay where all components are added simultaneously. Pre-incubation of mAbs in the absence of ACE2 was found to significantly overestimate the neutralizing capacity of the mAbs (Figure 3D), conferring the mAb a competitive advantage by allowing it to neutralize the RBD in the absence of competing ACE2. Although similarities in the overall trend of neutralization compared to the WT emerge (Supp Figure 4), the much lower relative IC_50_ values obtained in comparison to the competitive assay performed in parallel (Figure 3D), appreciably reduces the resolution of the differences in the neutralizing response between variants, in particular to N501Y, to which all mAbs demonstrated an attenuated neutralizing response in the competitive assay (Figure 3B). This observation was however not as apparent in the non-competitive assay (Figure 3D).

**Figure 4.**
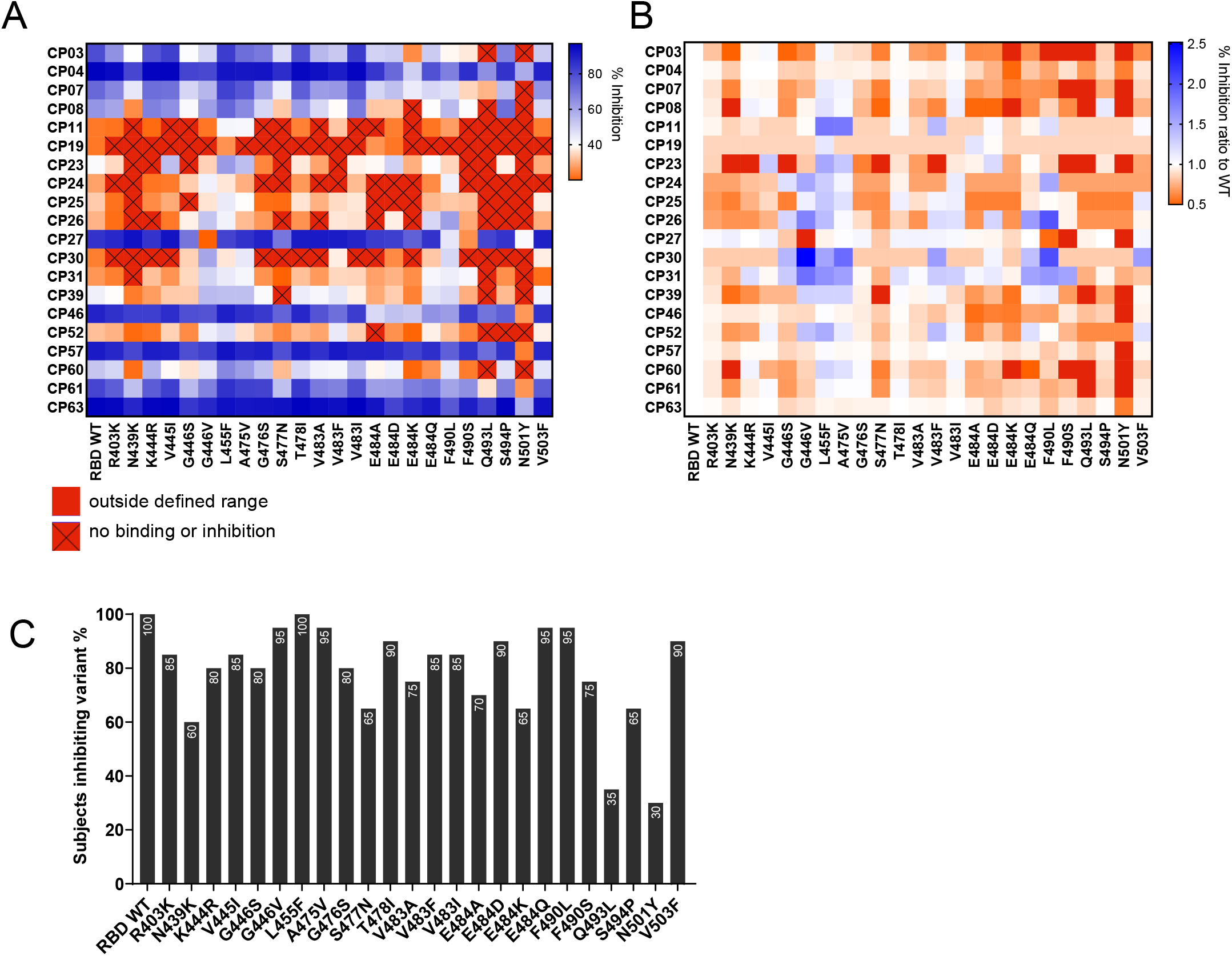
Mapping mutations to the RBD that affect inhibition of ACE-RBD by polyclonal antibodies. **A**. Percentage ACE2-RBD inhibition of a panel of *n*=20 convalescent samples (1 in 100 plasma dilution), demonstrating inhibition of RBD WT-ACE2 binding, mapped to the array of RBD variants in the multiplex assay. **B**. Percentage ACE2 Inhibition ratio to the WT for each subject to each RBD variant. **C**. Percentage of subjects which were able to block (inhibition >20%) binding to each RBD variant in the array. Data represents the mean of replicates.

### Mapping mutations to the SARS-CoV-2 RBD that affect recognition by convalescent human plasma antibodies

Having established the array of RBD natural variants and tested these with a panel of mAbs, we sought to evaluate recognition of the variants by polyclonal immune plasma from 20 individuals who had recovered from SARS-CoV-2 infection a mean of 36 days earlier (Table S1). These samples were collected early in the pandemic (March-April 2020) before the detection of the N501Y variant in Australia.

Plasma samples were tested at a single-point dilution of 1 in 100 across the array of natural RBD variants, based on the previously strong correlation of this dilution with relative IC_50_ values (Figure 1G). As observed in Figure 4A, most subjects demonstrated a reduction in the inhibition of most RBD variants, with the neutralizing response to variant N501Y being the most significantly attenuated, with many subjects demonstrating a ratio of less than 0.5 to that observed with the WT (Figure 4B), equating to over a 2-fold decrease in % inhibition to variant N501Y compared to the WT. Similar to the findings observed with the panel of mAbs, many convalescent plasma samples were able to bind the N501Y mutant with similar affinity to WT (Supplementary Figure 5), however likely due to the enhanced affinity of N501Y to ACE2, polyclonal convalescent plasma demonstrated a reduced ability to inhibit the ACE2-RBD interaction (Figure 4A and 4B) to this variant. Figure 4C illustrates the % of subjects which had a neutralizing response (i.e. >20%) to each RBD variant, with the lowest proportion of responders observed to occur to high affinity variants N501Y (30%) and Q493L (35%). This was followed by N439K (60%), N477N (65%), and E484K (65%). These results suggest that despite having the capacity to inhibit WT RBD, many convalescent individuals may not generate antibodies with high enough affinity capable of competing with RBD variants with an enhanced affinity to ACE2.

## Discussion

To date numerous SARS-CoV-2 RBD variant viruses have emerged, with the number and frequency of these variants steadily increasing since the beginning of the pandemic. Of further concern has been the observation that a number of these variants demonstrate escape from antibody neutralization (23-27,46), with the more recent emergence of new variants with multiple mutations in the RBD (6, 8), also demonstrating resistance to *in vitro* antibody neutralization (47). In the context of increasing population level immunity to WT SARS-CoV-2, and the rollout of vaccines and antibody therapies targeting the RBD, mapping the antibody response to emerging RBD mutations will be critical to guide future preparedness efforts by enabling pre-emptive forecasting of mutations that might impact antibody recognition.

To address the need to rapidly survey the neutralizing capacity of antibodies to RBD variants, here we describe a novel rapid high-throughput assay to measure RBD binding and neutralizing antibodies to an array of RBD variants. Using a panel of plasma samples obtained from subjects previously infected with SARS-CoV-2, we found this assay correlated well with a cytopathic effect virus microneutralization assay. Though this assay is not intended to replace gold standard SARS-CoV-2 cell-based neutralization assays, it provides a simple solution for broadly surveying the diversity of SARS-CoV-2 neutralizing specificities to an array of RBD mutants in a rapid high-throughput format, which can easily adapt to include new RBD variants as they emerge. Furthermore, this assay presents several key advantages over existing assays, since there is no need for viruses, cells, biosafety containment requirements, or highly skilled tissue-culture operators. Finally, results can be obtained rapidly on the same day in a high-throughput manner with the use of minimal sample volumes.

It has now been reported by several studies (23-27, 46) that antibody neutralization to some RBD mutants is reduced, suggesting that SARS-CoV-2 has mutated to evade host immunity. Profiling of a subset of previously published mAbs and polyclonal convalescent plasma samples, we demonstrate the heterogeneity of the neutralizing response to an array of RBD variants. Specifically, within the mAb panel we found variants at position 446 and 484 (with the exception of E484D) were poorly neutralized. We also found variant G446V which has demonstrated resistance to REGEN10987 (48) and C135 (27) to be poorly neutralized by our panel of mAbs. Furthermore F490L, which has been described to be remarkably resistant to mAbs (46) demonstrated weak binding to the panel of mAbs by comparison to the other variants in the array, with RBD alanine mutation at this position confirming that this is a critical epitope for certain mAbs. Despite this, we found most mAbs were able to demonstrate a modest inhibition at position 490. It is important to note that that residues L455, A475, F490 and Q493, are residues which have been previously reported to be SARS-CoV-2 RBD-ACE2 interacting residues based on structure analysis (12, 49). Unlike the previously described variants, which demonstrated escape from mAb recognition, N501Y was recognized at relatively high affinity by most mAbs, however inhibition to N501Y was attenuated across most mAbs, suggesting immune escape was likely due to the enhanced affinity of this variant for ACE2. We also observed via BLI an almost 5-fold greater affinity of the N501Y variant to ACE2 compared to WT, confirming previous studies that describe the formation of three additional bonds which increase the affinity of this variant to the ACE2 receptor (22, 42).

Although the E484K mutation has been associated with immune escape (27), predictions regarding its affinity to the ACE2 receptor are conflicting (22, 50). Our findings, which demonstrate a reduced affinity of E484K to ACE2 however support the recent observations in which the E484K mutation has been found to prevent the formation of two salt bridges that help to form and stabilize the RBD-ACE2 complex, thus reducing ACE2 binding affinity (22). We also found S477N (51), and Q493L, a variant at a position which interfaces directly with ACE2, had enhanced affinity to ACE2, confirming observations from deep mutational scanning analysis (42). What these differences in affinity to the ACE2 receptor mean for the overall viral fitness and transmissibility of these variants is beginning to be explored (52). However, lessons learned from the original SARS-CoV outbreak, which have served to guide SARS-COV-2 research suggest that the infectivity of different SARS-CoV strains in host cells is correlated to the binding affinity between the RBD of each strain and ACE2 (53-55). Indeed, the frequency and presence of the N501Y mutation in recently rapidly emerging SARS-CoV-2 linages, B.1.1.7, B.1.351, B.1.1.70 and P.1 appears to further strengthen this hypothesis, as does the frequency of S477N and S494P which we find bind ACE2 with an enhanced affinity (Figure 2B and 2D). It is interesting to note, however, that unlike higher affinity N501Y and S477N variants, F490L and G446V demonstrated the weakest EC_50_ values in our ACE2 binding assay (Figure 2D), but are observed at relatively high frequencies (Figure 2B), supporting the hypothesis that modestly deleterious mutations may have the potential to rise in prevalence if they confer escape from selective pressures such as the immune response (27, 56). In this array, we compared single amino acid RBD variants, however future studies aim to expand the array to incorporate the multiple amino acid mutations present in the RBD of newly emerging linages, as well as other variants which have in time come to rise in frequency.

When we profiled the polyclonal response of SARS-CoV-2 convalescent plasma to the RBD variants in array, we observed a similar pattern in the attenuation of the inhibitory response observed with the mAb panel to variants at position E484 and N501. This suggests that the majority of subjects infected early in the pandemic, despite having the capacity to inhibit RBD WT, have a limited repertoire of antibodies capable of competing with RBD variants with higher affinity to ACE2. This observation was especially pronounced for variants N501Y and Q493L, followed by N439K, and S477N. Alternatively, certain variants such as E484K may attenuate the neutralizing activity of convalescent plasma because it is situated in an immunodominant epitope targeted by most infected subjects. Though our findings are concordant with a number of previous studies (23-25, 27, 46, 52) it is still unknown what impact attenuation of the neutralization response to these variants is likely to have upon re-infection and vaccine efficacy, and what degree of neutralization is needed to sustain protection. The present study also observed improved neutralization in our assay when antibody was pre-incubated first, in the absence of the ACE2 receptor (Figure 3D), which raises to question whether current assays which employ similar methods may be overestimating the neutralizing response, particularly to high affinity RBD variants, such as N501Y. When the two assay formats were compared, we found a significant reduction in relative IC_50_ values when mAbs were allowed to neutralize the RBD prior to incubation with ACE2. These findings suggest that a competitive format where both mAbs and ACE2 (or ACE2 expressing cells) are added simultaneously, may allow for a more physiologically relevant assessment of the neutralizing capacity of NAbs. This approach may be particularly important when assessing mAbs therapeutics in infected individuals, or in immunocompromised vaccinated individuals where low levels of NAbs are present.

As SARS-CoV-2 continues to infect people globally, and variants with multiple mutations in the spike protein emerge, there is a growing urgency to rapidly develop effective and therapeutic options especially for high-risk individuals who may not develop a robust immune response or those that may not be able to be vaccinated. This novel high throughput multiplex assay has the potential to rapidly flag variants that may potentially demonstrate an enhanced affinity to ACE2. It also characterises the RBD neutralizing response in a competitive format, which takes into account the relative affinities of each RBD variant to the ACE2 receptor, to more accurately reflect the physiological dynamics of infection. The application of this novel assay thus covers a major gap in surveying and anticipating patterns of antibody resistance to emerging SARS-CoV-2 RBD, a factor which is becoming increasingly important for guiding the development of effective SARS-CoV-2 therapeutic agents to curb the ongoing pandemic.

## METHODS

### Materials Availability

This study did not generate new unique reagents.

### Data and code Availability

The raw datasets supporting the current study are available from the corresponding author on request.

### Ethics statement

The study protocols were approved by the University of Melbourne Human Research Ethics Committee and all associated procedures were carried out in accordance with approved guidelines. All participants provided written informed consent in accordance with the Declaration of Helsinki.

## EXPERIMENTAL SAMPLES AND SUBJECT DETAILS

### Plasma

Patients who had recovered from COVID-19 and healthy controls were recruited through contacts with the investigators and were invited to provide a blood sample as previously described (38). For all participants, whole blood was collected with sodium heparin anticoagulant, and plasma was collected and stored at -80 °C until use.

### Recombinant proteins

DNA encoding the truncated human ACE2 ectodomain (residues 19–613) with a C-terminal AVI-tag and 6xHis-tag was synthesised (IDT) and cloned into a pHLSec expression plasmid. Plasmid DNA was used for transient expression in Expi293F cells using Expifectamine 293 transfection kits (Thermofisher Scientific). Expression supernatant was harvested at day 6, and dialysed into 10mM Tris pH 8.0. ACE2 was subsequently purified using weak anion exchange (DEAE Sepharose, Cytiva), followed by size exclusion chromatography (Superdex 200, Cytiva). Protein was then biotinylated enzymatically using BirA enzyme and further purified using strong anion exchange (MonoQ, Cytiva).

Human IgG1 Anti-SARS-CoV-2 RBD Neutralizing Antibody, (#SAD-S35, Acro Biosystems) and mouse IgG2b SARS-CoV-2 RBD Neutralizing Antibody (#40592-MM43, Sino Biologicals) were used as positive controls. An in-house influenza mAb and human monoclonal CR3022 which binds RBD at an epitope that does not overlap with the ACE2 binding site of SARS-CoV-2 and therefore shows no competition with ACE2 (41), were used as negative controls. The heavy and light chain sequences of previously described monoclonal antibodies COVA2-15 and COVA-18 (43), and C002 and C135 (44) were synthesised and cloned into human IgG1 expression plasmids. These were expressed in Expi293 cells and purified using Protein A.

### RBD natural variant multiplex array

For the natural variant array 24 SARS-CoV-2 RBD variants were selected from the GISAID RBD surveillance repository (June 2020) for inclusion in the array. The WHU1 wild type (WT) isolate (2), SARS-CoV-2 RBD sequence (GenBank: MN908947.3. (amino acids 319–541; RVQP…CVNF), with the signal peptide (amino acids 1–14; MFVF…VSSQ) and a hexahistidine tag cloned into pcDNA3.4 vectors by Genscript Corporation (Piscataway, NJ, USA) was used as the original WT sequence. SARS-CoV-2 Spike S1 (#40591-V08H, Sino Biologicals) recombinant protein was also included in the array. RBD variants were expressed in Expi293 HEK cells (ThermoFisher), maintained in suspension at 37 °C and 8% CO_2_. Cells were transfected at a density of 3 x 10^6^ with 1 µg of plasmid DNA for 1 mL of culture and ExpiFectamine™ 293 reagent diluted in Opti-Mem™ (ThermoFisher) following the manufacturer’ s protocol. 22 hours after transfection, ExpiFectamine™ 293 transfection Enhancer 1 and 2 (ThermoFisher) was added to transfected cells along with lupin peptone. Six days after transfection, the supernatant was collected by centrifugation, and 10 mM MgCl_2_ was added to improve binding to the column and filtered through a 0.22 µm filter. RBD variants were purified by loading the supernatant onto a 1 mL Ni Excel column (GE Healthcare). Columns were equilibrated and washed using Dulbecco’ s phosphate buffered saline (DPBS). RBD variants were eluted using 300 mM imidazole, 100 mM NaCl DPBS buffer. A second purification step was performed by loading the eluate on a S75 increase 10/300 pg gel filtration column (GE Healthcare). Protein concentration was determined by absorbance measurement at 280 nm and purity was determined using SDS-PAGE.

Alanine mutants were generated by Genscript Corporation (Piscataway, NJ, USA). Where an alanine was naturally present, a serine substation was used instead. Recombinant proteins were expressed from HEK293 cells and subsequently purified using Ni-NTA columns. Protein concentration was determined by absorbance measurement at 280 nm and purity was determined using SDS-PAGE.

RBD variant multiplex bead cocktails were generated as previously described (57) Briefly, each respective RBD protein was coupled to a distinct magnetic carboxylated bead region (Bio Rad) using a two-step carbodiimide reaction at a ratio of 1 million beads to 5 µg of each RBD protein. RBD multiplex assays were conducted in black, clear bottom 384-well plates (Greiner Bio-One). 20µl of RBD natural variant cocktail (700 beads of each bead region per well) were added to 10 µl of mAbs at a starting final concentration of 80nM per well, prepared as 8-point 4-fold titrations in assay buffer (0.1% BSA/PBS). For the ACE2 inhibition assay 20µl of 25µg/ml of AviTagged Biotinylated ACE2 was subsequently added to all wells, whereas 20µl of assay buffer was added to RBD antibody binding assay wells, to bring the total final volume per well to 50µl. Each plate was incubated for 2 hours on a plate shaker at RT, before being washed twice in 0.05% PBS Tween 20. For the ACE2 inhibition assay, binding was detected with 40 µl of Streptavidin, R-Phycoerythrin Conjugate (SAPE) (#S866, Thermo Fisher) at 4µg/ml added for 1 hour, followed by the addition of 10 µl of 10 µg/ml of R-Phycoerythrin Biotin-XX Conjugate (#P811, Thermo Fisher), incubating for an additional hour. Relative RBD antibody binding was detected using anti-human IgG R-Phycoerythrin (PE) Conjugate (#9040-09, Southern Biotech) at 1.3µg/ml for 2 hours. For both assays’ plates were incubated on a plate shaker at RT before being washed three times. 60 µl of sheath buffer was added to each well, with plates left to shake for 10 minutes on a plate shaker prior to acquisition on a FlexMap3D(tm) (Luminex Corporation). The binding of ACE2 or IgG detected as phycoerythrin-labelled reporter is measured as MFI (Median Fluorescence Intensity). Coupling efficiency of the recombinant RBD was assessed as above using the anti-His Tag antibody (#A00174, GenScript), in place of the IgG detector conjugate.

The non-competitive ACE2 inhibition assay was performed as above with the exception that antibody was allowed to incubate first for 1 hour with the RBD variant bead cocktail, before addition of biotinylated ACE2 for a further hour.

### Microneutralization test

The virus microneutralization test was performed as previously described (38, 39). Briefly SARS-CoV-2 isolate CoV/Australia/VIC01/2020 (58) was passaged in Vero cells, and samples were serially diluted before the addition of 100 TCID_50_.of SARS-CoV-2 in MEM/0.5% BSA and incubation at room temperature for 1 h. Residual virus infectivity in the plasma/virus mixtures was assessed in quadruplicate wells of Vero cells incubated in serum-free media containing 1 μg ml^-1^ of TPCK trypsin at 37 °C and 5% CO_2_ viral cytopathic effect was read on day 5.

### Biolayer Interferometry (BLI)

The affinity and kinetic constants of selected RBD variants to the ACE-2 receptor were measured by BLI performed on the Octet Red instrument (FortéBio). Assays were performed in black 96 well plates at 30°C with agitation at 1000rpm. Streptavidin Sensors (FortéBio) were hydrated for 20 minutes in kinetic buffer 0.01 M HEPES, 0.15 M NaCl, 3 mM EDTA, 0.005% v/v Surfactant P20, pH 7.4 (GE healthcare), prior to loading of 3µg/ml of biotinylated ACE2 ligand for 180s. Following loading, the baseline signal was recorded for 120s in kinetic buffer. Sensors were then immersed into wells containing 2-fold serial dilutions of each recombinant SARS-CoV-2 RBD variant in kinetic buffer for 150 s (association phase) starting at 100nM, this was followed by immersion in kinetic buffer for 360s (dissociation phase). Curve fitting was performed using a global fit 1:1 binding model using Octet Data Analysis software v12.0.2.3 (FortéBio), and baseline drift was corrected by reference subtracting the shift of an ACE2 loaded sensor immersed in kinetic buffer only. Mean kinetic constant values from two independent experiments were determined, with all binding curves matching the theoretical fit with an r^2^ value of > 0.99. The complex half-time (t½) in seconds was calculated using the formula: t½ = In2/k_dis_=∼0.69/ k_dis_.

## QUANTIFICATION AND STATISTICAL ANALYSIS

All statistical and non-linear regression analysis was performed with GraphPad Prism v9.0. The arithmetic mean of replicate neutralization measurements and the arithmetic mean of replicate measurements in other assays were used in the correlation (Spearman) and regression analyses for other measurements. Maximal ACE2 binding MFI was determined by the mean (quadruplicate) buffer only i.e. ACE2 only (no inhibitor) controls. % ACE2 binding inhibition was calculated as ACE2 Inhibition (%) = (1 – Sample ACE2 MFI/Maximal ACE2 MFI) x100. All data generated from multiplex assays represent the mean of values repeated independently twice. Throughout the manuscript, significance was defined as *p* < 0.05.

## Data Availability

All relevant data is available in the manuscript or in the supplementary materials.

## Acknowledgments

We would thank Kathleen Wragg, Robyn Esterbauer, Helen E. Kent and Thakshila Amarasena (University of Melbourne) for their assistance.

This study was supported by the Victorian Government and Medical Research Future Fund (MRFF) GNT2002073 (to W.-H.T., D.I.G., A.W.C., S.J.K. and A.K.W.), Emergent Ventures Fast Grant (A.W.C.) and the Paul Ramsay Foundation (D.I.G., S.J.K., A.K.W and A.W.C). D.I.G., W.-H.T., S.J.K., A.K.W. and A.W.C are supported by NHMRC fellowships. W-H.T. is a Howard Hughes Medical Institute–Wellcome Trust International Research Scholar (208693/Z/17/Z). This work was also supported in part by funding from the Jack Ma Foundation (D.I.G, D.F.J.P, A.K.W, K.S, A.W.C). and the A2 Milk Company (KS). KS is supported by an NHMRC Investigator grant. N.A.G. is supported by an ARC DECRA fellowship. The Melbourne WHO Collaborating Centre for Reference and Research on Influenza is supported by the Australian Government Department of Health.

The authors acknowledge the Victorian State Government Operational Infrastructure Support and Australian Government NHMRC IRIISS.

## Authors contributions

E.L and A.W.C drafted the original manuscript. E.L, E.R.H, K.S. and A.W.C. designed experimental protocols. E.L, E.R.H, A.A, M.O.N, F.L.M, K.J.S, and S.D performed experiments. A.K.W, P.P, J.A.J., S.R, N.A.G., L.H., D.F.J.P, D.I.G., W-H.T contributed unique reagents. S.J.K and J.A.J. contributed unique samples. E.L. and A.W.C. analysed data. E.L and E.R.H generated final figures.

A.W.C. conceived and supervised the study. All authors reviewed the manuscript.

## Competing interests

The authors declare no competing interests.

## Data and materials availability

All relevant data is available in the manuscript or in the supplementary materials. Any additional supporting data can be provided upon request.

## Supplementary figures

**Supp Figure 1. A**. Determination of the ACE2-RBD inhibition cut-off of 20% based of testing *n=*12 SARS-CoV-2 negative samples at a 1 in 100 plasma dilution, with the light dotted line representing 1 standard deviation from the mean, and the solid dotted line representing the assay cut-off at the Mean + 2 Standard Deviations **B**. Pearson’ s correlation and linear regression between the mean % inhibition obtained from testing convalescent SARS-CoV-2 samples on two independent days, by two operators **C**. Spearman’ s correlation of the mean % ACE2 inhibition obtained from two independent multiplex runs vs the microneutralization titer obtained for each sample

**Supp Figure 2. Coupling Efficiency of RBD variants**

Coupling efficiency of each recombinant RBD variant was measured on the multiplex assay with 10µg/ml of anti-His Tag antibody. Coupling of each variant was compared to the WT RBD. All included variants were coupled to at least 75% relative to WT as indicated by the dotted line. Variants with a calculated >100% coupling efficiency to WT are capped to 100%. Bars represent mean± SD, determined from two independent experiments.

**Supp Figure 3. Single-plex vs Multi-Plex Comparison**

MFI values of wells containing a single bead region coupled to an RBD variant were assayed and compared to MFI values obtained for the full array of RBD variants in a single well (multi-plex). Data represent the mean of quadruplicates. **A**. Linear regression between IgG Single-plex vs Multi-plex MFI **B**. Linear regression between ACE2 inhibition Single-plex vs Multi-plex MFI

**Supp Figure 4. Non-Competitive assay values of RBD natural variant ACE2-RBD IC**_**50**_. **inhibition relative to RBD WT**.

Blue -Stronger inhibition of RBD variants relative to WT (IC_50_. < RBD WT) Orange-Weaker inhibition of RBD variants relative to WT (>1-6 fold RBD WT IC_50_.). Red >6 fold weaker RBD WT IC_50_. (Very poor/absence of inhibition)

**Supp Figure 5. Heat Map of Mean IgG Binding (MFI) of polyclonal convalescent plasma samples to each respective RBD variant**.

Blue – High level of IgG Binding to RBD variant Orange-Weak Binding, Outside defined range = Red-Very Low Level of IgG Binding RBD variant (MFI <5,000). No binding to RBD Variant (MFI <1,000)

## Notes

### Competing Interest Statement

The authors have declared no competing interest.

### Author Declarations

The study protocols were approved by the University of Melbourne Human Research Ethics Committee

